# 1^st^, 2^nd^, and 3^rd^+ SARS-CoV-2 infections: associations of prior infections with protection and severity

**DOI:** 10.1101/2025.04.07.25324191

**Authors:** Hannah E. Maier, Sergio Ojeda, Abigail Shotwell, Roger Lopez, Nery Sanchez, Saira Saborio, Miguel Plazaola, Carlos Barilla, Guillermina Kuan, Angel Balmaseda, Aubree Gordon

## Abstract

Since the emergence of the antigenically distinct Omicron variant, SARS-CoV-2 reinfections have increased and are common. Yet, as we approach endemicity, the frequency, severity, and consequences of reinfections remain poorly understood. Using data from a household transmission study in Managua, Nicaragua (2020-2024), we evaluated protection conferred by one, two, and three+ prior infections and compared the severity of first, second, and third+ RT-PCR-confirmed SARS-CoV-2 infections. In adults, compared to those with no prior infections, after adjusting for vaccination, one, two, and three+ prior infections were associated with increasing protection from symptomatic infection: 46%, (95% Confidence Interval [CI]: 32-57%), 69% (95%CI: 56-78%), and 77% (95%CI: 58-88%), respectively. Compared to first infections, second and third+ infections were associated with decreasing severity in adults, adjusted for vaccination; 35% (95%CI: 11-52%) and 42% (95%CI: 6-65%) less moderate/severe disease, and 152% (95%CI: 46-336%) and 243% (95%CI: 82-545%) more subclinical disease, respectively.

---

It is still unclear what endemic SARS-CoV-2 will look like and whether future infections and disease will differ from today. Population immunity continues to build through repeated infection and vaccination, yet the virus keeps evolving, complicating efforts to achieve protection. In this work, we use data from a community-based prospective cohort study in Nicaragua, a setting with high infection and vaccination rates that is likely close to a stable level of endemicity. We measure the protection associated with prior infection by comparing SARS-CoV-2 incidence among individuals with one, two, and three+ prior infections, and we compare the severity of first, second, and third+ SARS-CoV-2 infections.

Between March 2020 and December 2023, 2,389 people aged 0-95 years participated in the Household Influenza Cohort Study (HICS) contributing a total of 9,397 person-years (PY) of follow-up. Sex distribution was balanced among children, but adult male participation was lower (Supplementary Figure 2). In Nicaragua, the original SARS-CoV-2 strain circulated in 2020, Gamma and Delta predominated in 2021, and Omicron became dominant starting in 2022^1,2^.

ELISA completeness was high, with blood samples available for an average of 95% of enrolled participants during sampling times (Supplementary Table 1).

We detected a total of 3,651 serologically- or RT-PCR-confirmed infections among HICS participants. Over the course of their enrollment, 1,050 participants (44.0%) had one detected infection, 788 (33.0%) two infections, 324 (13.6%) three+ infections, and 227 (9.5%) had no detected infections. The majority of those remaining uninfected either left the cohort in 2020/2021 (n=162, 71.4%) or were enrolled as infants (n=32, 14.1%). By October 2021, 91.9% of HICS participants had been infected (93.4% of adults), and by October 2024, 97.6% had been infected (99.7% of adults, Figure 1A).

**Figure 1.**
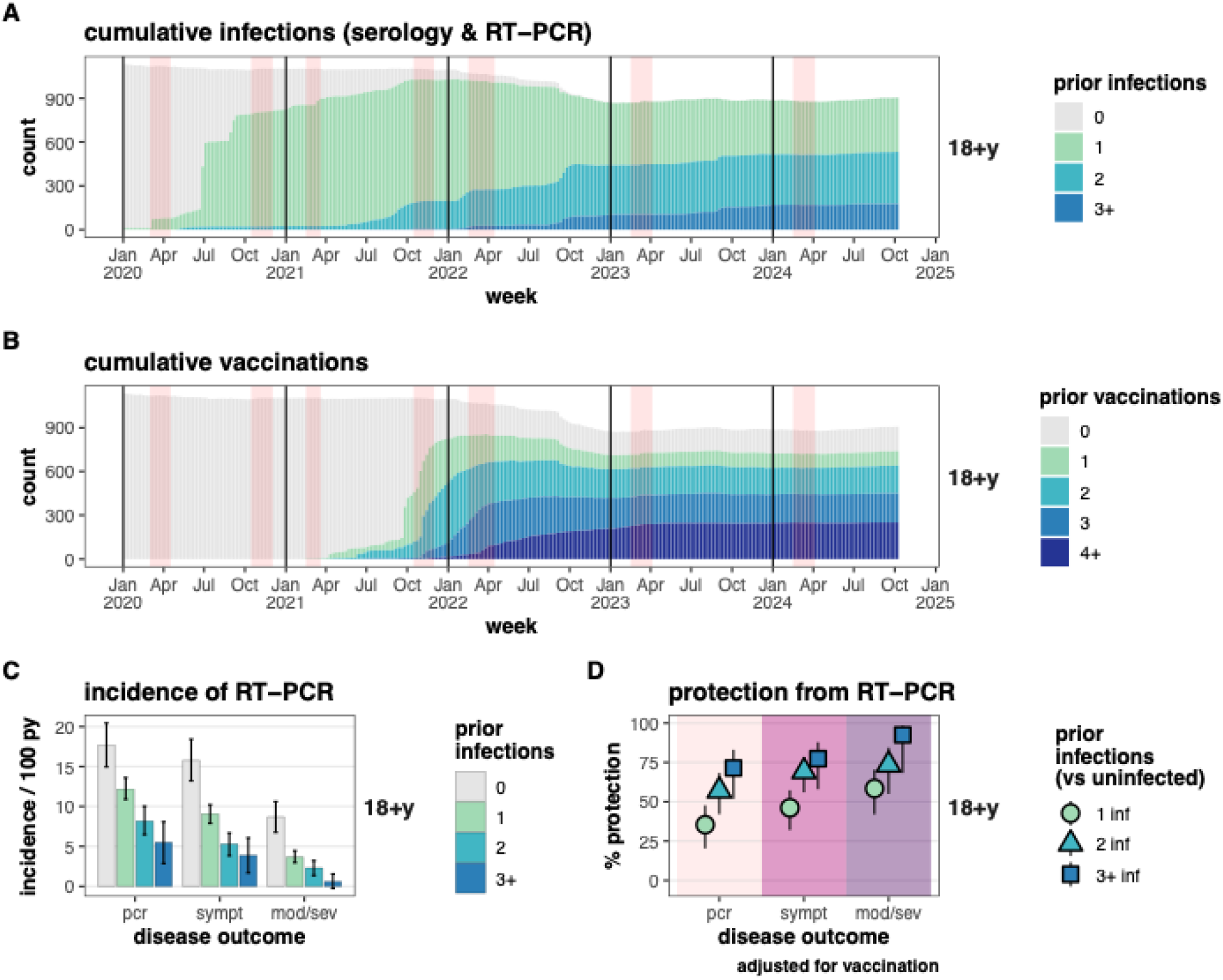
Protection from SARS-CoV-2 infection associated with prior infections in adults. A) Cumulative infections and B) cumulative vaccinations among adults. Vertical red bands indicate the timing of annual and midyear blood samples. C) Incidence of and D) Protection (1 – risk ratio) against RT-PCR-detected SARS-CoV-2 infection, with outcomes of all RT-PCR-detected infections (pcr), symptomatic RT-PCR infections (sympt), and moderate or severe RT-PCR infections (mod/sev). PY = person-years.

Of the 3,651 total infections, 1,146 (31.4%) were detected by RT-PCR. Only 2 RT-PCR-detected infections were missing any symptom data; these 2 participants were excluded from analyses. Among the 1,144 RT-PCR-confirmed infections with symptom data, 346 were first, 614 were second, and 184 were third+ infections (Figure 2A&B).

**Figure 2.**
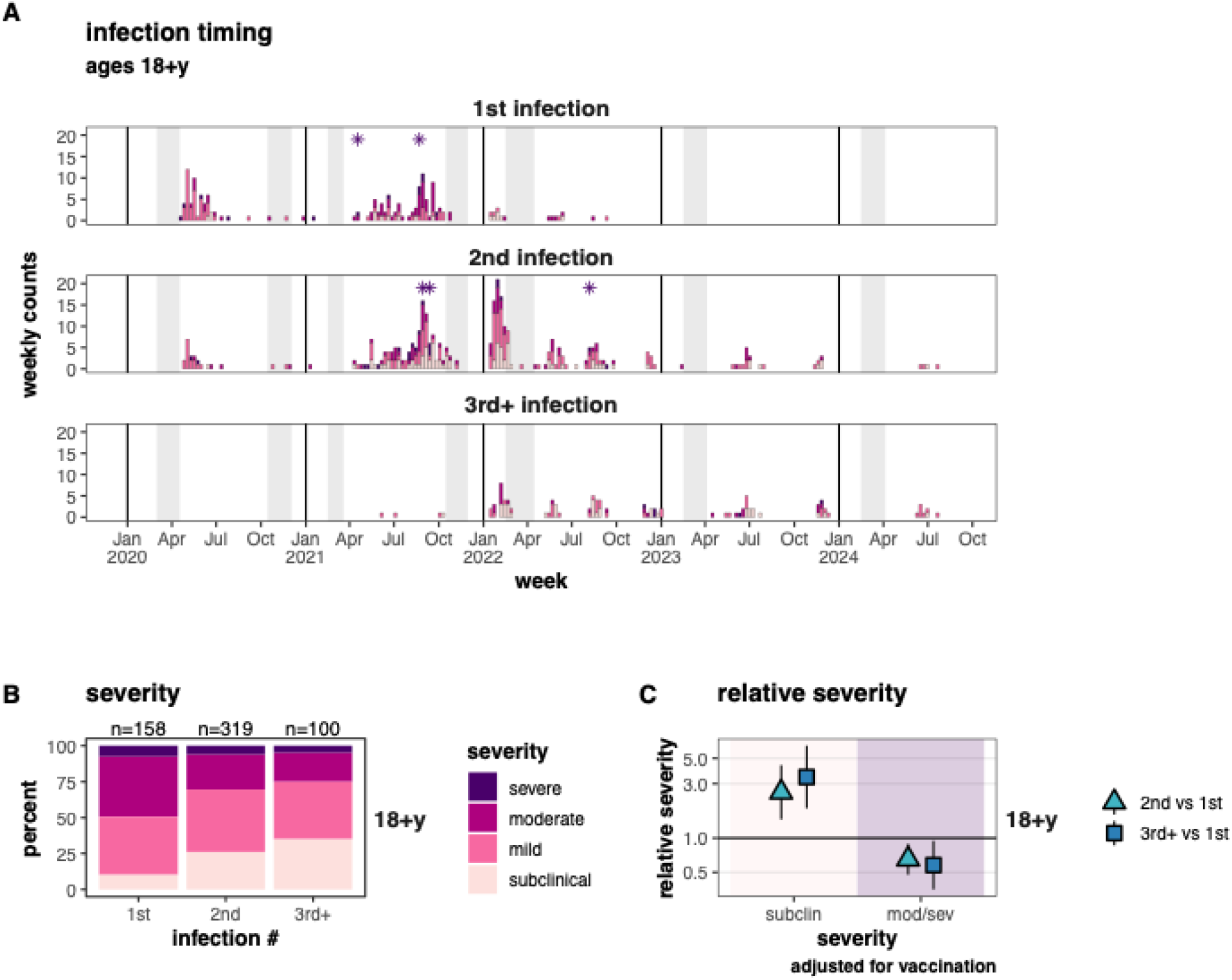
Severity of 1st, 2nd, and 3rd+ SARS-CoV-2 infections in adults. A) Timing of 1st, 2nd, and 3rd+infections, filled in by severity. Asterisks indicate SARS-CoV-2-related deaths. Vertical grey bands indicate the timing of annual and midyear blood samples. B) severity of 1st, 2nd, and 3rd+ infections, and C) relative severity comparing 2nd and 3rd+ infections to 1st infections.

Nicaragua did not have access to vaccines as early as many other parts of the world. When vaccines did become more widely available around September-October 2021, they included a broader selection of lesser-studied SARS-CoV-2 vaccines ^3^. A large-scale vaccination effort followed, and by January 2022, 70.0% of HICS participants had been vaccinated (78.3% of adults, Figure 1B). We adjusted for vaccination and stratified by period to account for vaccination.

In adults, protection from reinfection increased with the number of prior infections and with the severity of the outcome (all RT-PCR-confirmed, symptomatic, and moderate/severe infections). The highest protection—92% (95%CI: 68-98%)—was observed against moderate/severe infections in those with three+ prior infections, while the lowest protection—35% (95%CI: 20-48%)—was against all RT-PCR-confirmed infections in those with one prior infection versus uninfected individuals (Figure 1 & Supplemental Table 2).

The incidence of symptomatic infections in adults was 15.8, 9.1, 5.3, and 3.9 cases per 100 PY for those with 0, 1, 2, and 3+ prior infections (Figure 1C), corresponding to 46%, (95%CI: 32-57%), 69% (95%CI: 56-78%), and 77% (95%CI: 58-88%) protection for those with 1, 2, and 3+ versus no prior infections (Figure 1D). For moderate/severe disease, incidence was 8.7, 3.7, 2.3, and 0.6 cases per 100 PY in those with 0, 1, 2, and 3+ prior infections, corresponding to 58% (95%CI: 42-70%), 73% (95%CI: 55-84%), and 92% (95%CI: 68-98%) protection for those with 1, 2, and 3+ versus 0 prior infections.

Similarly, infections became increasingly less severe with each subsequent reinfection in adults (Figure 2 & Supplemental Table 3). The proportion of moderate/severe disease decreased from 49% to 30% to 25% in first, second, and third+ infections (Figure 2B), corresponding to reductions of 35% (95%CI: 11-52%) and 42% (95%CI: 6-65%) for second and third+ versus first infections (Figure 2C). Meanwhile, the proportion of subclinical infections increased from 10% to 26% to 35% in first, second, and third+ infections, corresponding to increases of 152% (95%CI: 46-336%) and 243% (95%CI: 82-545%) for second and third+ infections versus first infections.

Stratifying by variant era (Supplemental Figure 3A), we again observed increasing protection with each subsequent reinfection in adults, with higher protection in the pre-Omicron period (Supplemental Figure 3B & Supplemental Table 3). For example, adults with two prior infections in the pre-Omicron era had 92% protection (95%CI: 75-98%) against symptomatic infection, whereas during the Omicron era protection dropped to 80% (95%CI: 61-89%). However, in the Omicron era, more reinfections had occurred, and adults with three+ prior infections had 86% protection (95%CI: 67-94%) against symptomatic infection and 95% protection (95%CI: 77-99%) against moderate/severe infection.

Likewise, reductions in disease severity were more pronounced in the pre-Omicron period. During this time, second infections in adults resulted in 45% (95%CI: 20-63%) less moderate/severe disease and 275% (95%CI: 68-739%) more subclinical infections versus first infections (Supplemental Figure 4 & Supplemental Table 3). While comparing the Omicron to the pre-Omicron periods, adults had more subclinical infections (29% vs 22%) and less moderate/severe disease (26% vs 48%). However, during the Omicron era, there was little difference in severity by infection number.

Additionally, the period-of- and time-since-last infection were associated with protection from reinfection, and, to a lesser extent, severity (Supplemental Figure 5). As expected, protection was highest when the prior infection matched the circulating variant, and was more recent. The highest protection in adults was observed in the pre-Omicron era, with 98% (95%CI: 90-99%) protection against symptomatic infection for those with a prior pre-Omicron infection, and 98% (95%CI: 89-100%) protection from moderate/severe infection for those whose last infection occurred 0-180 days prior. Protection remained present even for adults with original SARS-CoV-2 infections and infections ≥1 year prior. For example, during the pre-Omicron era, protection against symptomatic infection was 75% (95%CI: 66-82%) for adults with an original SARS-CoV-2 infection, and 61% (95%CI: 44-72%) for those with an infection 1-2 years prior.

Children also had some protection (Supplemental Figure 6), and older children had a higher proportion of subclinical reinfections (Supplemental Figure 7). However, confidence intervals were much wider, and patterns were less consistent.

## DISCUSSION

Overall, we found that 1, 2, and 3+ prior infections were associated with increasing protection from and decreasing severity of reinfections. Protection was stronger pre-Omicron. Children had weaker protection, maybe because of smaller numbers, or because children’s immune responses to SARS-CoV-2 are different than adults’^4^.

Numerous studies have investigated protection from prior SARS-CoV-2 infection^1,5-10^, including hybrid immunity^5,6^, and have similarly reported lower protection against Omicron. Our study stands out among current research, because we have complete infection histories of participants, and our study population has complex infection and vaccination histories. Our estimates of protection against Omicron (all above 65%) were higher than many others, which reported protection around 40-45%^9,10^. One study found that having two prior infections provided no additional benefit^11^. While several studies found reduced severity for SARS-CoV-2 reinfections^12-14^, one reported increased severity for Omicron among those with prior infection ^15^. Some studies have been misinterpreted as suggesting reinfections are more severe^16^, but we found no studies specifically assessing severity across multiple reinfections. The ongoing decline in COVID-19 hospitalizations and deaths further supports the idea that reinfection severity decreases over time^17,18^.

The lack of reduced severity for reinfections during the Omicron era may reflect Omicron’s antigenic distance or indicate a limit to the protection conferred by immunity. Alternatively, reinfections during Omicron may indeed be less severe, but the difference may be most pronounced at the more severe end of the spectrum, which our study did not capture.

A major strength of this study is the HICS design—a long-running, population-based prospective cohort that spans the entire SARS-CoV-2 pandemic. These results are broadly representative of the general population and the inclusion of blood sampling allowed for the detection of asymptomatic SARS-CoV-2 infections, enabling accurate infection histories. Additionally, loss to follow-up was minimal, and symptom data was available for nearly all participants. However, sample sizes were small when stratifying by age and period, and we were unable to further stratify by Omicron subvariants. Additionally, we could not assess protection or severity reduction from bivalent or updated vaccines due to lack of availability.

In this highly-infected and vaccinated population, we observed reductions in both incidence and severity following multiple SARS-CoV-2 infections. However, during the Omicron era, reinfections did not appear to have reduced severity. These findings suggest that SARS-CoV-2 immunity continues developing, but this population may be approaching endemicity.

## Data Availability

All data used in the present study are available upon reasonable request to the Aubree Gordon.

## ACKNOWLEDGEMENTS

This work was supported by the National Institute of Allergy and Infectious Diseases at the National Institute of Health (award no. R01 AI120997 to A.G. and contract nos.

HHSN272201400006C and 75N93021C00016 to A.G.) ELISA reagents were funded by Open Philanthropy and the Michigan Center for Infectious Disease Threats. Aubree Gordon is supported by the Biosciences Initiative at the University of Michigan.

## METHODS

### Ethics Statement

This study was approved by the institutional review boards at the Nicaraguan Ministry of Health and the University of Michigan (HUM00119145 and HUM00178355). Informed consent or parental permission was obtained for all participants. Assent was obtained from children aged ≥6 years.

### The Household Influenza Cohort Study (HICS)

HICS is an ongoing household cohort study with an embedded household transmission study, located in district II of Managua, Nicaragua. It was started in 2017 to study influenza, and in 2020 SARS-CoV-2 was added. Regular blood samples are collected for all cohort participants, twice yearly in 2020 and 2021, then yearly thereafter. The transmission study is activated when a household member becomes sick and tests positive for SARS-CoV-2 by RT-PCR after seeking care at the study health clinic. Activated households are visited by study staff regularly, at approximately days 0, 3, 7, 14, 21, and 30, for collection of respiratory samples and symptom diaries.

### Laboratory Assays

Blood samples were tested as paired serum samples (current vs baseline) using an enzyme-linked immunosorbent assays (ELISAs) protocol adapted from the Krammer laboratory ^19^. The SARS-CoV-2 spike receptor binding domain (RBD), spike, and nucleoprotein (NP) proteins for ELISAs were produced in single batches at the Life Sciences Institute at the University of Michigan; these were generated based on the original SARS-CoV-2 strain. RBD was used for screening (positive/negative) as it is more specific than spike, and spike was used to titer samples that screened positive by RBD ELISA; NP ELISAs were also run, to distinguish between vaccine and infection responses. Real-time reverse-transcription polymerase chain reaction (RT-PCR) was performed according to the protocol from Chu et al. ^20^.

All RT-PCR and most ELISAs were performed at the Nicaraguan National Virology Laboratory, with a minority of 2020 annual samples processed at the University of Michigan.

### Infection histories

We used positive serology screens, antibody titer values, 4-fold rises in titer, and RT-PCR positivity to define first and secondary infections, as outlined in Supplemental Figure 1. To be defined as an infection, serological samples must have been from participants at least 6 months of age, to avoid incorrectly classifying maternal immunity, and first infections serologically detected by RBD screen or spike titer must have been before the first COVID vaccination.

Distinct infections detected serologically must have been ≥6 months apart from each other. Secondary infections detected serologically with spike must not have samples surrounding vaccination. RT-PCR-detected infections must have been ≥60 days apart from each other. Sequential infections detected by RT-PCR and 4-fold rise must have occurred >90 days apart from each other. Any potential infections within 60 days of each other were flagged for manual inspection. Only RT-PCR-confirmed infections were selected to compare severity.

### Severity

Symptoms reported in daily symptom diaries and clinic visits were used to define severity, as severe (hospitalized, doctors say they should have been hospitalized, or death), moderate (lower respiratory symptoms or overall poor condition), mild (loss of smell/taste, fever, or ≥2 symptoms), or subclinical (≤1 symptom not meeting definitions for higher severity).

### Analysis

For the protection analysis, person time was counted while participants were enrolled in HICS (some exited and re-entered, and the time not enrolled was not counted). While new households were recruited in 2023 to maintain the cohort size, for the protection analysis, participants were limited to those enrolled before January 1, 2021 or were born into the cohort (enrolled before age 1y) so that full infection histories would be known.

Poisson regression was used to calculate relative risks (RR). Percent protection was calculated as (1-RR)^*^100. Analyses were adjusted for vaccination status, defined as unvaccinated, partially vaccinated, fully vaccinated (number of doses varied by vaccine; Sputnik light only required 1 dose for full vaccination), or boosted; we counted vaccines that occurred at least 14 days prior. Vaccines targeting the SARS-CoV-2 nucleocapsid were not counted with the vaccinated groups (a small number of people have had a vaccine targeting the nucleocapsid, starting in 2023). We stratified all analyses by age (0-9y, 10-17y, and 18+y). We also stratified by variant era, to account for high vaccination rates starting around November 2021 and differences in variants.

## Code Availability

R code used for these analyses is available at https://github.com/hannahma/SARS-CoV-2_severity_1st_2nd_3rd

