## Supplement for "1^st^, 2^nd^, and 3^rd^+ SARS-CoV-2 infections: associations of prior infections with protection and severity"

Table of contents:

Figures:

Tables:

---

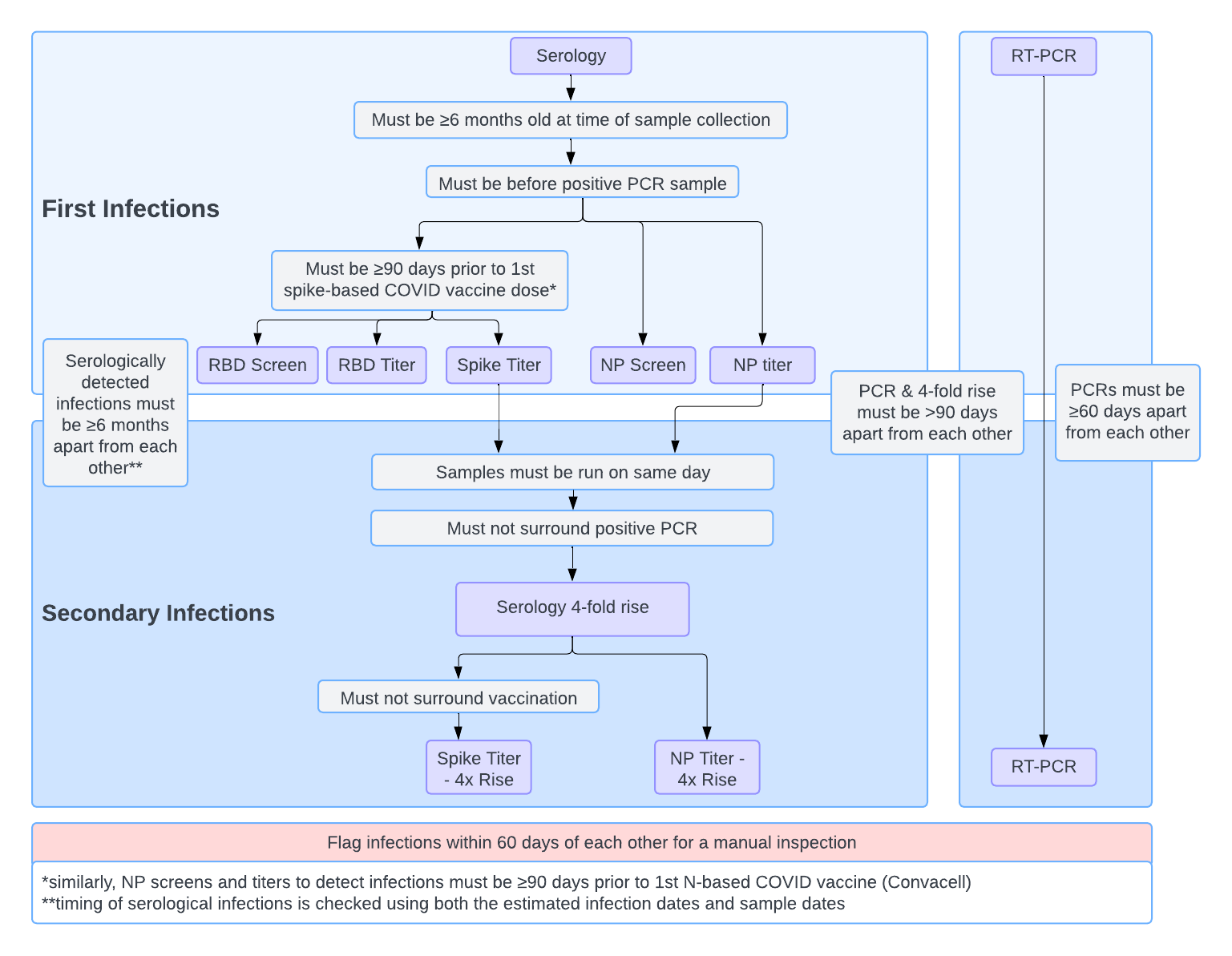

Figure S 1. Defining infection histories.

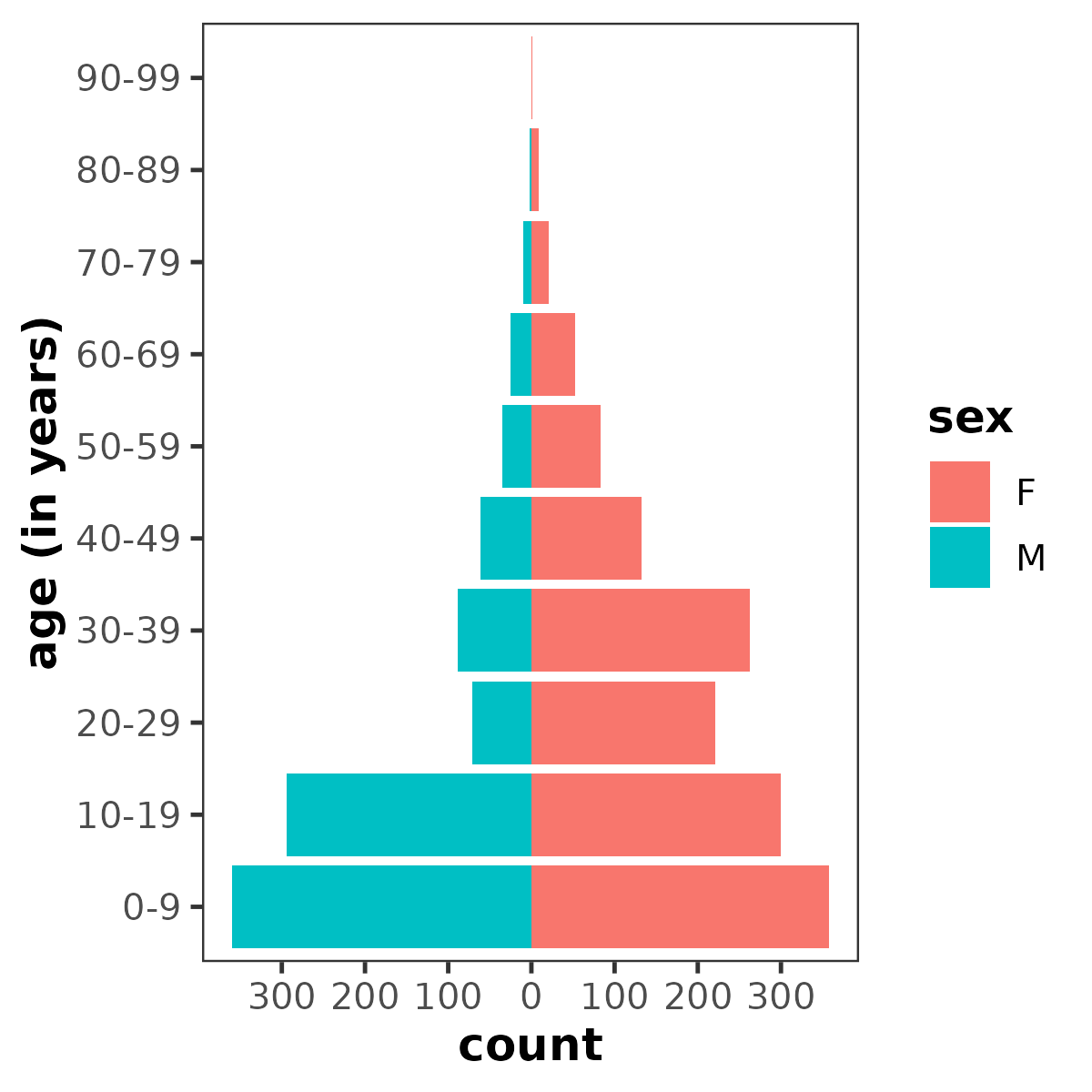

Figure S 2. HICS population pyramid.

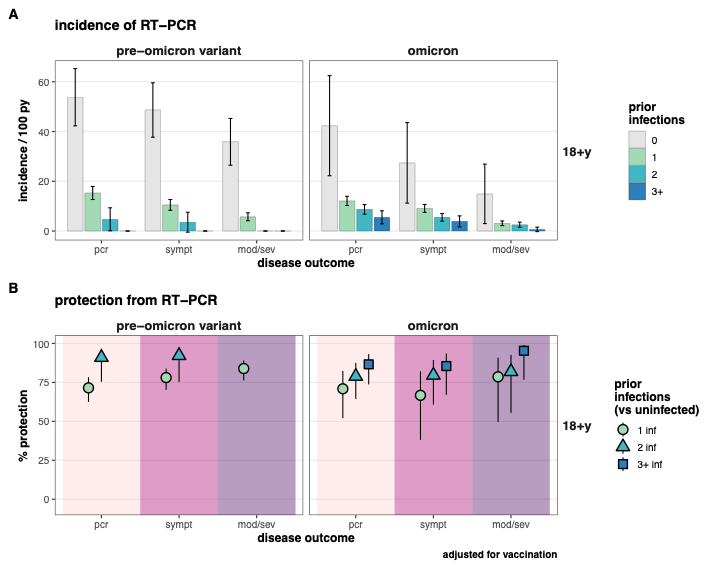

Figure S 3. Protection from SARS-CoV-2 infection associated with prior infections in adults, by period.

A) Incidence of and B) Protection (1 – risk ratio) against RT-PCR-detected SARS-CoV-2 infection, with outcomes of all RT-PCR-detected infections (pcr), symptomatic RT-PCR infections (sympt), and moderate or severe RT-PCR infections (mod/sev). PY = person-years.

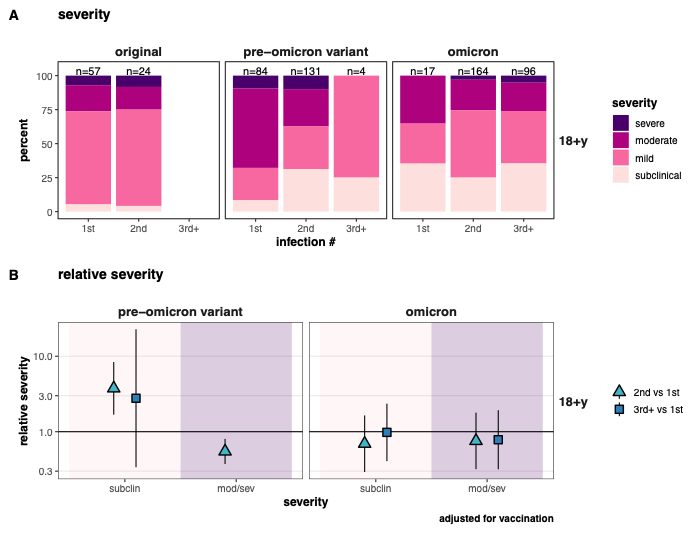

Figure S 4. Severity of 1st, 2nd, and 3rd+ SARS-CoV-2 infections in adults, by period.

A) severity of 1st, 2nd, and 3rd+ infections, and B) relative severity comparing 2nd and 3rd+ infections to 1st infections.

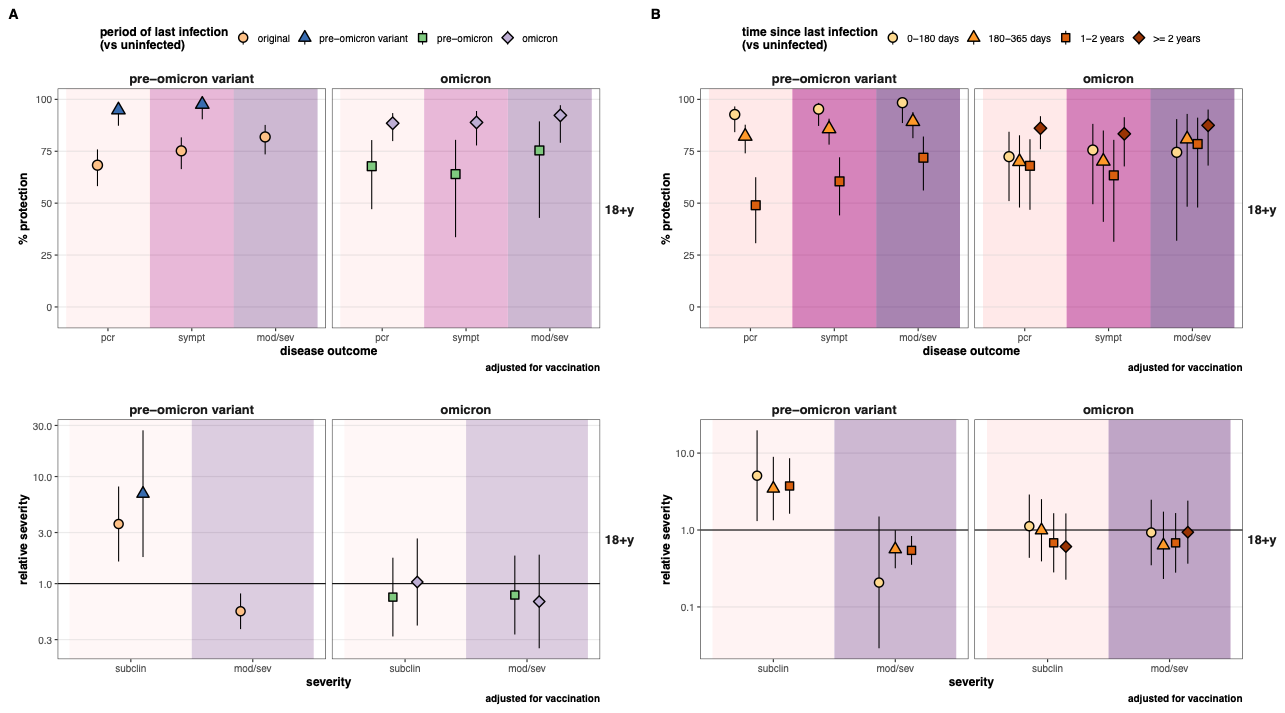

Figure S 5. Protection and severity associated with period of and time since last infection.

*A) period of last infection, and B) time since last infection. Top plots in A) and B) show protection (1 – risk ratio) against RT-PCR-detected SARS-CoV-2 infection, with outcomes of all RT-PCR-detected infections (pcr), symptomatic RT-PCR infections (sympt), and moderate or severe RT-PCR infections (mod/sev). Bottom plots show relative severities.*

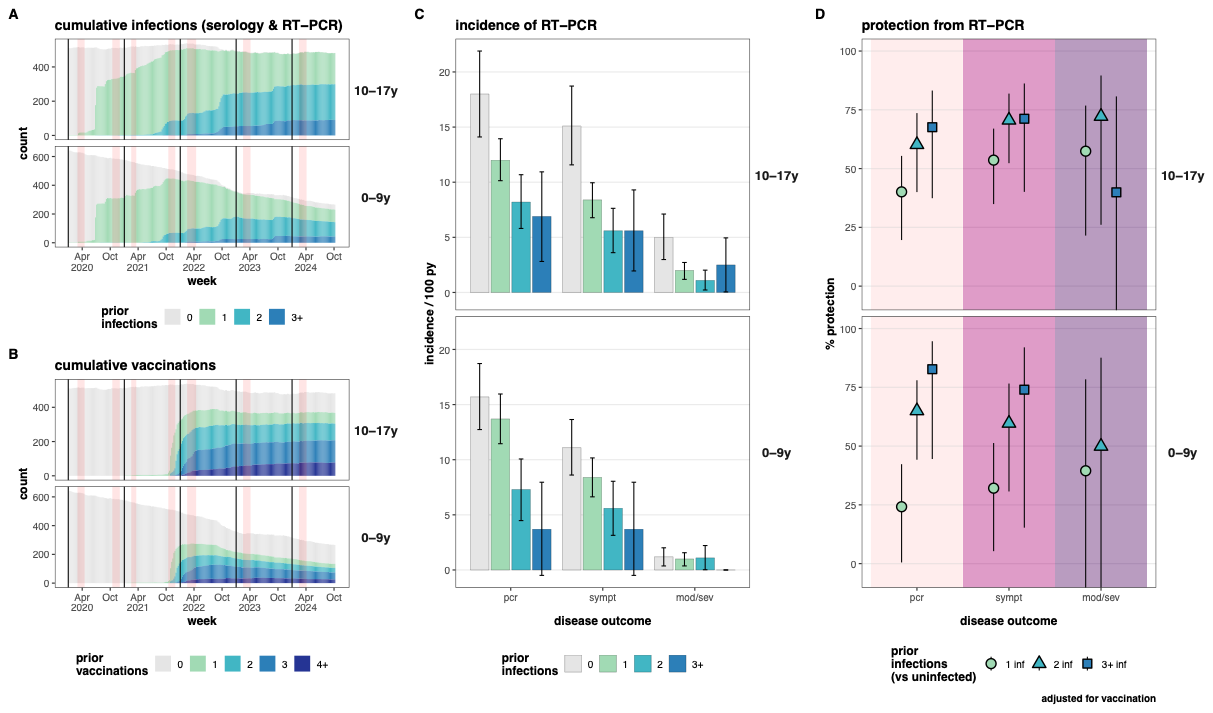

Figure S 6. Protection from SARS-CoV-2 infection associated with prior infections in kids.

A) Cumulative infections and B) cumulative vaccinations among kids. Vertical red bands indicate the timing of annual and midyear blood samples. C) Incidence of and D) Protection (1 – risk ratio) against RT-PCR-detected SARS-CoV-2 infection, with outcomes of all RT-PCR-detected infections (pcr), symptomatic RT-PCR infections (sympt), and moderate or severe RT-PCR infections (mod/sev). PY = person-years.

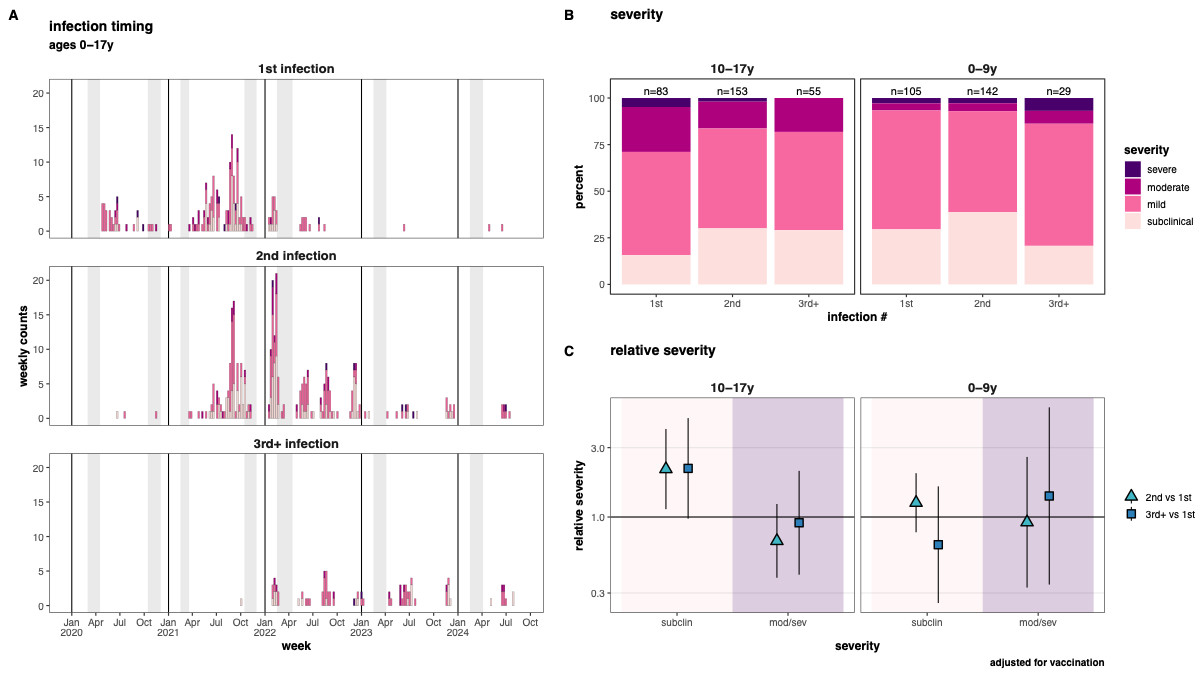

Figure S 7. Severity of 1st, 2nd, and 3rd+ SARS-CoV-2 infections in kids.

*A) Timing of 1st, 2nd, and 3rd+infections, filled in by severity. Vertical grey bands indicate the timing of annual and midyear blood samples. B) severity of 1st, 2nd, and 3rd+ infections, and C) relative severity comparing 2nd and 3rd+ infections to 1st infections.*

Table S 1 Serology samples and ELISA results

|  |  | 2020 | | 2021 | | 2022 | 2023 | 2024 |  |  |
| --- | --- | --- | --- | --- | --- | --- | --- | --- | --- | --- |
|  |  | annual | midyear | annual | midyear | annual | annual | annual | AVERAGE | TOTAL |
| SAMPLES | |  |  |  |  |  |  |  |  |  |
|  | # of people enrolled* | 2271 | 2206 | 2187 | 2162 | 2093 | 1804 | 1985 | 2101.1 | 2389 |
|  | # of samples (%**) | 2185 (96.2%) | 2021 (91.6%) | 2116 (96.7%) | 1863 (86.2%) | 1985 (94.8%) | 1788 (99.1%) | 1963 (98.9%) | 1998.7 (95.1%) | 13921 |
| SAMPLE RESULTS: n (%***) | |  |  |  |  |  |  |  |  |  |
|  | Spike/RBD | 2000 (91.5%) | 2019 (99.9%) | 2094 (99.0%) | 1863 (100.0%) | 1964 (98.9%) | 1663 (93.0%) | 5 (0.3%) | 1658.3 (83.4%) | 11608 (83.4%) |
|  | Nucleocapsid | 48 (2.2%) | 10 (0.5%) | 1567 (74.1%) | 1664 (89.3%) | 1904 (95.9%) | 1698 (95.0%) | 1572 (80.1%) | 1209.0 (60.8%) | 8463 (60.8%) |
| POSITIVITY: n (%****) | |  |  |  |  |  |  |  |  |  |
|  | Spike/RBD | 120 (6.0%) | 1135 (56.2%) | 1290 (61.6%) | 1760 (94.5%) | 1930 (98.3%) | 1663 (100.0%) | 5 (100%) | 1129 (68.1%) | 7903 (68.1%) |
|  | Nucleocapsid | 6 (12.5%) | 3 (30.0%) | 862 (55.0%) | 1330 (79.9%) | 1715 (90.1%) | 1643 (96.8%) | 1500 (95.4%) | 1008.4 (83.4%) | 7059 (83.4%) |
| * | at time of sample |  |  |  |  |  |  |  |  |  |
| ** | people with samples / people enrolled | | | |  |  |  |  |  |  |
| *** | samples with results / samples collected | | | |  |  |  |  |  |  |
| **** | positive samples (by screen or titer) / samples with results | | |  |  |  |  |  |  |  |

Table S 2 Protection from SARS-CoV-2 reinfection

|  | # prior  infections |  |  |  | # of RT-PCR-confirmed  infections | | |  | Incidence | | |  | Incidence Rate Ratio* | | |
| --- | --- | --- | --- | --- | --- | --- | --- | --- | --- | --- | --- | --- | --- | --- | --- |
| subset |  | PY | n |  | all | sympt | mod/sev |  | all | sympt | mod/sev |  | all | sympt | mod/sev |
| all | 0 | 2027.5 | 2355 |  | 347 | 286 | 109 |  | 17.1 | 14.1 | 5.4 |  | ref | ref | ref |
|  | 1 | 4921.0 | 2162 |  | 615 | 430 | 132 |  | 12.5 | 8.7 | 2.7 |  | 0.66 (0.57, 0.77) | 0.56 (0.47, 0.66) | 0.46 (0.35, 0.62) |
|  | 2 | 1898.2 | 1112 |  | 153 | 103 | 33 |  | 8.1 | 5.4 | 1.7 |  | 0.41 (0.33, 0.51) | 0.33 (0.26, 0.43) | 0.30 (0.19, 0.46) |
|  | 3+ | 550.6 | 324 |  | 31 | 24 | 6 |  | 5.6 | 4.4 | 1.1 |  | 0.29 (0.20, 0.42) | 0.27 (0.17, 0.41) | 0.19 (0.08, 0.44) |
| 18+y | 0 | 897.9 | 1165 |  | 159 | 142 | 78 |  | 17.7 | 15.8 | 8.7 |  | ref | ref | ref |
|  | 1 | 2605.7 | 1192 |  | 319 | 236 | 97 |  | 12.2 | 9.1 | 3.7 |  | 0.65 (0.52, 0.80) | 0.54 (0.43, 0.68) | 0.42 (0.30, 0.58) |
|  | 2 | 1006.9 | 608 |  | 83 | 53 | 23 |  | 8.2 | 5.3 | 2.3 |  | 0.43 (0.32, 0.58) | 0.31 (0.22, 0.44) | 0.27 (0.16, 0.45) |
|  | 3+ | 310.2 | 186 |  | 17 | 12 | 2 |  | 5.5 | 3.9 | 0.6 |  | 0.29 (0.17, 0.48) | 0.23 (0.12, 0.42) | 0.08 (0.02, 0.32) |
| 10-17y | 0 | 455.6 | 570 |  | 82 | 69 | 23 |  | 18.0 | 15.1 | 5.0 |  | ref | ref | ref |
|  | 1 | 1279.2 | 693 |  | 154 | 107 | 25 |  | 12.0 | 8.4 | 2.0 |  | 0.60 (0.45, 0.80) | 0.46 (0.33, 0.65) | 0.43 (0.23, 0.79) |
|  | 2 | 533.8 | 378 |  | 44 | 30 | 6 |  | 8.2 | 5.6 | 1.1 |  | 0.40 (0.26, 0.60) | 0.29 (0.18, 0.48) | 0.28 (0.10, 0.74) |
|  | 3+ | 160.0 | 110 |  | 11 | 9 | 4 |  | 6.9 | 5.6 | 2.5 |  | 0.32 (0.17, 0.63) | 0.29 (0.14, 0.60) | 0.60 (0.19, 1.87) |
| 0-9y | 0 | 674.0 | 707 |  | 106 | 75 | 8 |  | 15.7 | 11.1 | 1.2 |  | ref | ref | ref |
|  | 1 | 1036.1 | 575 |  | 142 | 87 | 10 |  | 13.7 | 8.4 | 1.0 |  | 0.76 (0.58, 1.00) | 0.68 (0.49, 0.95) | 0.61 (0.22, 1.69) |
|  | 2 | 357.6 | 255 |  | 26 | 20 | 4 |  | 7.3 | 5.6 | 1.1 |  | 0.35 (0.22, 0.56) | 0.40 (0.23, 0.69) | 0.50 (0.12, 2.02) |
|  | 3+ | 80.4 | 61 |  | 3 | 3 | 0 |  | 3.7 | 3.7 | 0.0 |  | 0.17 (0.05, 0.56) | 0.26 (0.08, 0.85) | not calculated |
| original | 0 | 1492.6 | 2315 |  | 94 | 87 | 23 |  | 6.3 | 5.8 | 1.5 |  | ref | ref | ref |
|  | 1 | 717.2 | 1488 |  | 27 | 25 | 6 |  | 3.8 | 3.5 | 0.8 |  | not calculated | not calculated | not calculated |
|  | 2 | 14.4 | 27 |  | 0 | 0 | 0 |  | 0.0 | 0.0 | 0.0 |  | not calculated | not calculated | not calculated |
| pre- | 0 | 392.5 | 712 |  | 205 | 164 | 78 |  | 52.2 | 41.8 | 19.9 |  | ref | ref | ref |
| omicron | 1 | 1626.6 | 1980 |  | 236 | 154 | 59 |  | 14.5 | 9.5 | 3.6 |  | 0.28 (0.23, 0.34) | 0.23 (0.19, 0.29) | 0.18 (0.13, 0.25) |
| variant | 2 | 136.6 | 354 |  | 5 | 3 | 0 |  | 3.7 | 2.2 | 0.0 |  | 0.07 (0.03, 0.18) | 0.06 (0.02, 0.18) | not calculated |
|  | 3+ | 2.6 | 8 |  | 0 | 0 | 0 |  | 0.0 | 0.0 | 0.0 |  | not calculated | not calculated | not calculated |
| omicron | 0 | 142.3 | 201 |  | 48 | 35 | 8 |  | 33.7 | 24.6 | 5.6 |  | ref | ref | ref |
|  | 1 | 2577.3 | 1753 |  | 352 | 251 | 67 |  | 13.7 | 9.7 | 2.6 |  | 0.40 (0.29, 0.54) | 0.39 (0.27, 0.56) | 0.47 (0.22, 0.99) |
|  | 2 | 1747.3 | 1102 |  | 148 | 100 | 33 |  | 8.5 | 5.7 | 1.9 |  | 0.25 (0.18, 0.35) | 0.23 (0.16, 0.34) | 0.35 (0.16, 0.76) |
|  | 3+ | 548.0 | 324 |  | 31 | 24 | 6 |  | 5.7 | 4.4 | 1.1 |  | 0.17 (0.11, 0.26) | 0.18 (0.11, 0.30) | 0.20 (0.07, 0.59) |
| *adjusted for vaccination (partial, full, boosted vs unvaccinated) | | | | | | | |  |  |  |  |  |  |  |  |

(continued on next page)

|  | # prior  infections |  |  |  | # of RT-PCR-confirmed  infections | | |  | Incidence | | |  | Incidence Rate Ratio* | | |
| --- | --- | --- | --- | --- | --- | --- | --- | --- | --- | --- | --- | --- | --- | --- | --- |
| subset |  | PY | n |  | all | sympt | mod/sev |  | all | sympt | mod/sev |  | all | sympt | mod/sev |
| 18+y, | 0 | 701.6 | 1156 |  | 58 | 55 | 16 |  | 8.3 | 7.8 | 2.3 |  | ref | ref | ref |
| original | 1 | 393.5 | 830 |  | 24 | 23 | 6 |  | 6.1 | 5.8 | 1.5 |  | not calculated | not calculated | not calculated |
|  | 2 | 13.3 | 24 |  | 0 | 0 | 0 |  | 0.0 | 0.0 | 0.0 |  | not calculated | not calculated | not calculated |
| 18+y, | 0 | 156.2 | 283 |  | 84 | 76 | 56 |  | 53.8 | 48.7 | 35.9 |  | ref | ref | ref |
| pre- | 1 | 857.2 | 1028 |  | 131 | 90 | 49 |  | 15.3 | 10.5 | 5.7 |  | 0.29 (0.22, 0.38) | 0.22 (0.16, 0.30) | 0.16 (0.11, 0.24) |
| omicron | 2 | 84.9 | 199 |  | 4 | 3 | 0 |  | 4.7 | 3.5 | 0.0 |  | 0.09 (0.03, 0.25) | 0.08 (0.02, 0.25) | not calculated |
| variant | 3+ | 1.7 | 5 |  | 0 | 0 | 0 |  | 0.0 | 0.0 | 0.0 |  | not calculated | not calculated | not calculated |
| 18+y, | 0 | 40.2 | 73 |  | 17 | 11 | 6 |  | 42.3 | 27.4 | 14.9 |  | ref | ref | ref |
| omicron | 1 | 1355.0 | 960 |  | 164 | 123 | 42 |  | 12.1 | 9.1 | 3.1 |  | 0.29 (0.18, 0.48) | 0.33 (0.18, 0.62) | 0.21 (0.09, 0.50) |
|  | 2 | 908.7 | 601 |  | 79 | 50 | 23 |  | 8.7 | 5.5 | 2.5 |  | 0.21 (0.12, 0.36) | 0.20 (0.11, 0.39) | 0.18 (0.07, 0.45) |
|  | 3+ | 308.5 | 186 |  | 17 | 12 | 2 |  | 5.5 | 3.9 | 0.6 |  | 0.13 (0.07, 0.26) | 0.15 (0.06, 0.33) | 0.05 (0.01, 0.23) |
| 10-17y, | 0 | 341.3 | 555 |  | 18 | 16 | 4 |  | 5.3 | 4.7 | 1.2 |  | ref | ref | ref |
| original | 1 | 168.3 | 364 |  | 2 | 1 | 0 |  | 1.2 | 0.6 | 0.0 |  | not calculated | not calculated | not calculated |
|  | 2 | 0.6 | 2 |  | 0 | 0 | 0 |  | 0.0 | 0.0 | 0.0 |  | not calculated | not calculated | not calculated |
| 10-17y, | 0 | 90.9 | 180 |  | 55 | 45 | 17 |  | 60.5 | 49.5 | 18.7 |  | ref | ref | ref |
| pre- | 1 | 403.2 | 533 |  | 57 | 36 | 8 |  | 14.1 | 8.9 | 2.0 |  | 0.24 (0.16, 0.35) | 0.18 (0.12, 0.28) | 0.10 (0.04, 0.24) |
| omicron | 2 | 28.6 | 88 |  | 0 | 0 | 0 |  | 0.0 | 0.0 | 0.0 |  | not calculated | not calculated | not calculated |
| variant | 3+ | 0.6 | 2 |  | 0 | 0 | 0 |  | 0.0 | 0.0 | 0.0 |  | not calculated | not calculated | not calculated |
| 10-17y, | 0 | 23.3 | 36 |  | 9 | 8 | 2 |  | 38.6 | 34.3 | 8.6 |  | ref | ref | ref |
| omicron | 1 | 707.7 | 549 |  | 95 | 70 | 17 |  | 13.4 | 9.9 | 2.4 |  | 0.32 (0.16, 0.63) | 0.26 (0.13, 0.55) | 0.26 (0.06, 1.11) |
|  | 2 | 504.5 | 374 |  | 44 | 30 | 6 |  | 8.7 | 5.9 | 1.2 |  | 0.21 (0.10, 0.43) | 0.16 (0.07, 0.36) | 0.13 (0.03, 0.62) |
|  | 3+ | 159.4 | 110 |  | 11 | 9 | 4 |  | 6.9 | 5.6 | 2.5 |  | 0.16 (0.07, 0.39) | 0.15 (0.06, 0.40) | 0.25 (0.05, 1.40) |
| 0-9y, | 0 | 449.8 | 670 |  | 18 | 16 | 3 |  | 4.0 | 3.6 | 0.7 |  | ref | ref | ref |
| original | 1 | 155.3 | 334 |  | 1 | 1 | 0 |  | 0.6 | 0.6 | 0.0 |  | not calculated | not calculated | not calculated |
|  | 2 | 0.4 | 1 |  | 0 | 0 | 0 |  | 0.0 | 0.0 | 0.0 |  | not calculated | not calculated | not calculated |
| 0-9y, | 0 | 145.4 | 264 |  | 66 | 43 | 5 |  | 45.4 | 29.6 | 3.4 |  | ref | ref | ref |
| pre- | 1 | 366.2 | 499 |  | 48 | 28 | 2 |  | 13.1 | 7.6 | 0.5 |  | 0.30 (0.21, 0.44) | 0.27 (0.17, 0.44) | 0.17 (0.03, 0.87) |
| omicron | 2 | 23.1 | 72 |  | 1 | 0 | 0 |  | 4.3 | 0.0 | 0.0 |  | 0.11 (0.02, 0.79) | not calculated | not calculated |
| variant | 3+ | 0.2 | 1 |  | 0 | 0 | 0 |  | 0.0 | 0.0 | 0.0 |  | not calculated | not calculated | not calculated |
| 0-9y, | 0 | 78.9 | 97 |  | 22 | 16 | 0 |  | 27.9 | 20.3 | 0.0 |  | ref | ref | ref |
| omicron | 1 | 514.6 | 420 |  | 93 | 58 | 8 |  | 18.1 | 11.3 | 1.6 |  | 0.60 (0.37, 0.96) | 0.54 (0.30, 0.95) | not calculated |
|  | 2 | 334.1 | 250 |  | 25 | 20 | 4 |  | 7.5 | 6.0 | 1.2 |  | 0.24 (0.14, 0.44) | 0.28 (0.14, 0.55) | not calculated |
|  | 3+ | 80.1 | 61 |  | 3 | 3 | 0 |  | 3.7 | 3.7 | 0.0 |  | 0.12 (0.04, 0.41) | 0.18 (0.05, 0.61) | not calculated |

*adjusted for vaccination (partial, full, boosted vs unvaccinated)

Table S 3 Severity of 1^st^, 2^nd^, and 3^rd^+ SARS-CoV-2 infections

|  | # prior  infections |  |  | # of RT-PCR-confirmed infections | | |  | Relative Severity* | |
| --- | --- | --- | --- | --- | --- | --- | --- | --- | --- |
| subset |  | n |  | subclinical | mild | mod/sev |  | subclinical | mod/sev |
| all | 1st | 346 |  | 60 (17.3%) | 177 (51.2%) | 109 (31.5%) |  | ref | ref |
|  | 2nd | 614 |  | 184 (30.0%) | 298 (48.5%) | 132 (21.5%) |  | 1.74 (1.29, 2.36) | 0.71 (0.55, 0.93) |
|  | 3rd+ | 184 |  | 57 (31.0%) | 88 (47.8%) | 39 (21.2%) |  | 1.82 (1.23, 2.70) | 0.74 (0.49, 1.10) |
| 18+y | 1st | 158 |  | 16 (10.1%) | 64 (40.5%) | 78 (49.4%) |  | ref | ref |
|  | 2nd | 319 |  | 83 (26.0%) | 139 (43.6%) | 97 (30.4%) |  | 2.52 (1.46, 4.36) | 0.65 (0.48, 0.89) |
|  | 3rd+ | 100 |  | 35 (35.0%) | 40 (40.0%) | 25 (25.0%) |  | 3.43 (1.82, 6.45) | 0.58 (0.35, 0.94) |
| 10-17y | 1st | 83 |  | 13 (15.7%) | 46 (55.4%) | 24 (28.9%) |  | ref | ref |
|  | 2nd | 153 |  | 46 (30.1%) | 82 (53.6%) | 25 (16.3%) |  | 2.14 (1.13, 4.03) | 0.68 (0.38, 1.23) |
|  | 3rd+ | 55 |  | 16 (29.1%) | 29 (52.7%) | 10 (18.2%) |  | 2.16 (0.97, 4.79) | 0.91 (0.40, 2.08) |
| 0-9y | 1st | 105 |  | 31 (29.5%) | 67 (63.8%) | 7 (6.7%) |  | ref | ref |
|  | 2nd | 142 |  | 55 (38.7%) | 77 (54.2%) | 10 (7.0%) |  | 1.25 (0.79, 2.00) | 0.92 (0.33, 2.59) |
|  | 3rd+ | 29 |  | 6 (20.7%) | 19 (65.5%) | 4 (13.8%) |  | 0.64 (0.26, 1.63) | 1.40 (0.34, 5.68) |
| original | 1st | 93 |  | 7 (7.5%) | 64 (68.8%) | 22 (23.7%) |  | ref | ref |
|  | 2nd | 27 |  | 2 (7.4%) | 19 (70.4%) | 6 (22.2%) |  | not calculated | not calculated |
| pre- | 1st | 205 |  | 40 (19.5%) | 86 (42.0%) | 79 (38.5%) |  | ref | ref |
| omicron | 2nd | 236 |  | 82 (34.7%) | 95 (40.3%) | 59 (25.0%) |  | 1.77 (1.21, 2.58) | 0.65 (0.46, 0.91) |
| variant | 3rd+ | 5 |  | 2 (40.0%) | 3 (60.0%) | 0 (0.0%) |  | 1.96 (0.47, 8.19) | not calculated |
| omicron | 1st | 48 |  | 13 (27.1%) | 27 (56.3%) | 8 (16.7%) |  | ref | ref |
|  | 2nd | 351 |  | 100 (28.5%) | 184 (52.4%) | 67 (19.1%) |  | 1.05 (0.59, 1.87) | 1.18 (0.57, 2.47) |
|  | 3rd+ | 179 |  | 55 (30.7%) | 85 (47.5%) | 39 (21.8%) |  | 1.12 (0.61, 2.07) | 1.36 (0.63, 2.93) |
| *adjusted for vaccination (partial, full, boosted vs unvaccinated) | | | | | |  |  |  |  |

(continued on next page)

|  | # prior  infections |  |  | # of RT-PCR-confirmed infections | | |  | Relative Severity* | |
| --- | --- | --- | --- | --- | --- | --- | --- | --- | --- |
| subset |  | n |  | subclinical | mild | mod/sev |  | subclinical | mod/sev |
| 18+y, | 1st | 57 |  | 3 (5.3%) | 39 (68.4%) | 15 (26.3%) |  | ref | ref |
| original | 2nd | 24 |  | 1 (4.2%) | 17 (70.8%) | 6 (25.0%) |  | not calculated | not calculated |
| 18+y, | 1st | 84 |  | 7 (8.3%) | 20 (23.8%) | 57 (67.9%) |  | ref | ref |
| pre-omicron | 2nd | 131 |  | 41 (31.3%) | 41 (31.3%) | 49 (37.4%) |  | 3.75 (1.68, 8.39) | 0.55 (0.37, 0.80) |
| variant | 3rd+ | 4 |  | 1 (25.0%) | 3 (75.0%) | 0 (0.0%) |  | 2.78 (0.34, 22.79) | not calculated |
| 18+y, | 1st | 17 |  | 6 (35.3%) | 5 (29.4%) | 6 (35.3%) |  | ref | ref |
| omicron | 2nd | 164 |  | 41 (25.0%) | 81 (49.4%) | 42 (25.6%) |  | 0.69 (0.29, 1.64) | 0.76 (0.32, 1.79) |
|  | 3rd+ | 96 |  | 34 (35.4%) | 37 (38.5%) | 25 (26.0%) |  | 0.98 (0.41, 2.36) | 0.78 (0.32, 1.93) |
| 10-17y, | 1st | 18 |  | 2 (11.1%) | 12 (66.7%) | 4 (22.2%) |  | ref | ref |
| original | 2nd | 2 |  | 1 (50.0%) | 1 (50.0%) | 0 (0.0%) |  | not calculated | not calculated |
| 10-17y, pre- | 1st | 56 |  | 10 (17.9%) | 28 (50.0%) | 18 (32.1%) |  | ref | ref |
| omicron variant | 2nd | 57 |  | 21 (36.8%) | 28 (49.1%) | 8 (14.0%) |  | 2.03 (0.95, 4.30) | 0.45 (0.20, 1.05) |
| 10-17y, | 1st | 9 |  | 1 (11.1%) | 6 (66.7%) | 2 (22.2%) |  | ref | ref |
| omicron | 2nd | 94 |  | 24 (25.5%) | 53 (56.4%) | 17 (18.1%) |  | 2.17 (0.29, 16.09) | 0.78 (0.18, 3.41) |
|  | 3rd+ | 55 |  | 16 (29.1%) | 29 (52.7%) | 10 (18.2%) |  | 2.35 (0.31, 17.89) | 0.77 (0.17, 3.60) |
| 0-9y, | 1st | 18 |  | 2 (11.1%) | 13 (72.2%) | 3 (16.7%) |  | ref | ref |
| original | 2nd | 1 |  | 0 (0.0%) | 1 (100.0%) | 0 (0.0%) |  | not calculated | not calculated |
| 0-9y, pre- | 1st | 65 |  | 23 (35.4%) | 38 (58.5%) | 4 (6.2%) |  | ref | ref |
| omicron | 2nd | 48 |  | 20 (41.7%) | 26 (54.2%) | 2 (4.2%) |  | 1.14 (0.62, 2.10) | 0.69 (0.13, 3.78) |
| variant | 3rd+ | 1 |  | 1 (100.0%) | 0 (0.0%) | 0 (0.0%) |  | 2.83 (0.38, 20.93) | not calculated |
| 0-9y, | 1st | 22 |  | 6 (27.3%) | 16 (72.7%) | 0 (0.0%) |  | ref | ref |
| omicron | 2nd | 93 |  | 35 (37.6%) | 50 (53.8%) | 8 (8.6%) |  | 1.30 (0.54, 3.12) | not calculated |
|  | 3rd+ | 28 |  | 5 (17.9%) | 19 (67.9%) | 4 (14.3%) |  | 0.60 (0.18, 2.00) | not calculated |
| *adjusted for vaccination (partial, full, boosted vs unvaccinated) | | | | | |  |  |  |  |
